# Development of a qualitative real-time RT-PCR assay for the detection of SARS-CoV-2: A guide and case study in setting up an emergency-use, laboratory-developed molecular assay

**DOI:** 10.1101/2020.08.26.20157297

**Authors:** Melis N. Anahtar, Bennett Shaw, Damien Slater, Elizabeth Byrne, Yolanda Botti-Lodovico, Gordon Adams, Stephen Schaffner, Jacqueline Eversley, Graham McGrath, Tasos Gogakos, Jochen Lennerz, Hetal Desai Marble, Lauren L. Ritterhouse, Julie Batten, N. Zeke Georgantas, Rebecca Pellerin, Sylvia Signorelli, Julia Thierauf, Molly Kemball, Christian Happi, Donald S. Grant, Daouda Ndiaye, Katherine J. Siddle, Samar B. Mehta, Jason Harris, Edward T. Ryan, Virginia Pierce, Regina LaRocque, Jacob E. Lemieux, Pardis Sabeti, Eric Rosenberg, John Branda, Sarah E. Turbett

## Abstract

Developing and deploying new diagnostic tests is difficult, but the need to do so in response to a rapidly emerging pandemic such as COVID-19 is crucially important for an effective response. In the early stages of a pandemic, laboratories play a key role in helping health care providers and public health authorities detect active infection, a task most commonly achieved using nucleic acid-based assays. While the landscape of diagnostics is rapidly evolving, polymerase chain reaction (PCR) remains the gold-standard of nucleic acid-based diagnostic assays, in part due to its reliability, flexibility, and wide deployment. To address a critical local shortage of testing capacity persisting during the COVID-19 outbreak, our hospital set up a molecular based laboratory developed test (LDT) to accurately and safely diagnose SARS-CoV-2. We describe here the process of developing an emergency-use LDT, in the hope that our experience will be useful to other laboratories in future outbreaks and will help to lower barriers to fast and accurate diagnostic testing in crisis conditions.

## BACKGROUND

In outbreak settings like that of COVID-19, there is an urgent need for rapid and reliable diagnostics that are widely deployable to identify infected individuals for medical care, to institute effective infection control measures, and to perform contact tracing. In normal, non-pandemic situations, our current system relies on large centralized diagnostic laboratories with specialized commercial equipment to perform diagnostic testing for infectious diseases. However, commercial diagnostics take time to develop, produce, and distribute, and are not widely available in a time of crisis. Simply put, laboratories and health care systems are often on their own to provide diagnostic testing until commercial assays become available, which can take weeks or months to develop and distribute. A solution to this problem is for laboratories to create LDTs, which can be done quickly and with supplies that are already on hand or easily available. In times of crisis, the regulatory standards that usually govern diagnostic testing are often relaxed, allowing laboratories much-needed flexibility. For example, in the United States the U.S. Food and Drug Administration’s Emergency Use Authorization (EUA) is intended to create a regulatory pathway to expand testing capacity and enable critical and rapid action in local laboratories to obviate the need for centralized testing.^1^ While regulation is an important factor and potential deterrent in the decision-making process of laboratories deciding to move forward with LDTs during a disease outbreak, technical hurdles are also crucially important. Even the most experienced and well-resourced laboratories can suffer from technical challenges that prevent them from developing a working diagnostic assay in a timely manner.

Within days of the first genome release of SARS-CoV-2, research groups from around the world began developing diagnostic assays. In the United States, for primarily regulatory reasons, as well as the tiered set-up of our Laboratory Response Network, that responsibility initially fell to the Centers for Disease Control and Prevention (CDC). However, by the time the U.S. began ramping up efforts to confront the pandemic in early March 2020, EUA-approved tests were still not widely available, commercially or otherwise. In response to the changing regulations and heightened demand for tests, clinical laboratories in the United States began developing assays and applying for their own EUAs.^2^ Our team, with support from both the state Department of Public Health (DPH) and long-standing collaborators, began to develop an LDT based on the CDC’s protocol on March 2, 2020 (Figure 1). We were then able to validate and implement a SARS-CoV-2 LDT for active clinical use by March 13, 2020, ahead of formal approval effective April 3, 2020.^3^ The LDT clinical assay served a vital role in diagnosing many of the early cases in our community, provided diagnoses in hospitalized patients, and allowed for enrollment of patients into impactful clinical trials. The demand for reliable diagnostic testing during the early stages of an outbreak is crucial, and therefore it is paramount that more clinical laboratories across the world feel empowered to perform their own testing to meet this need during future outbreaks. Most, if not all, clinical laboratories should be able to procure the materials and personnel to set up their own LDT and thus implement a nucleic-acid based diagnostic test within their own community. Both the CDC’s EUA and our LDT are based on standard molecular diagnostic principles, making this type of in-house protocol validation relatively straightforward and translatable as a roadmap for future testing of any pathogen.

**Figure 1.**
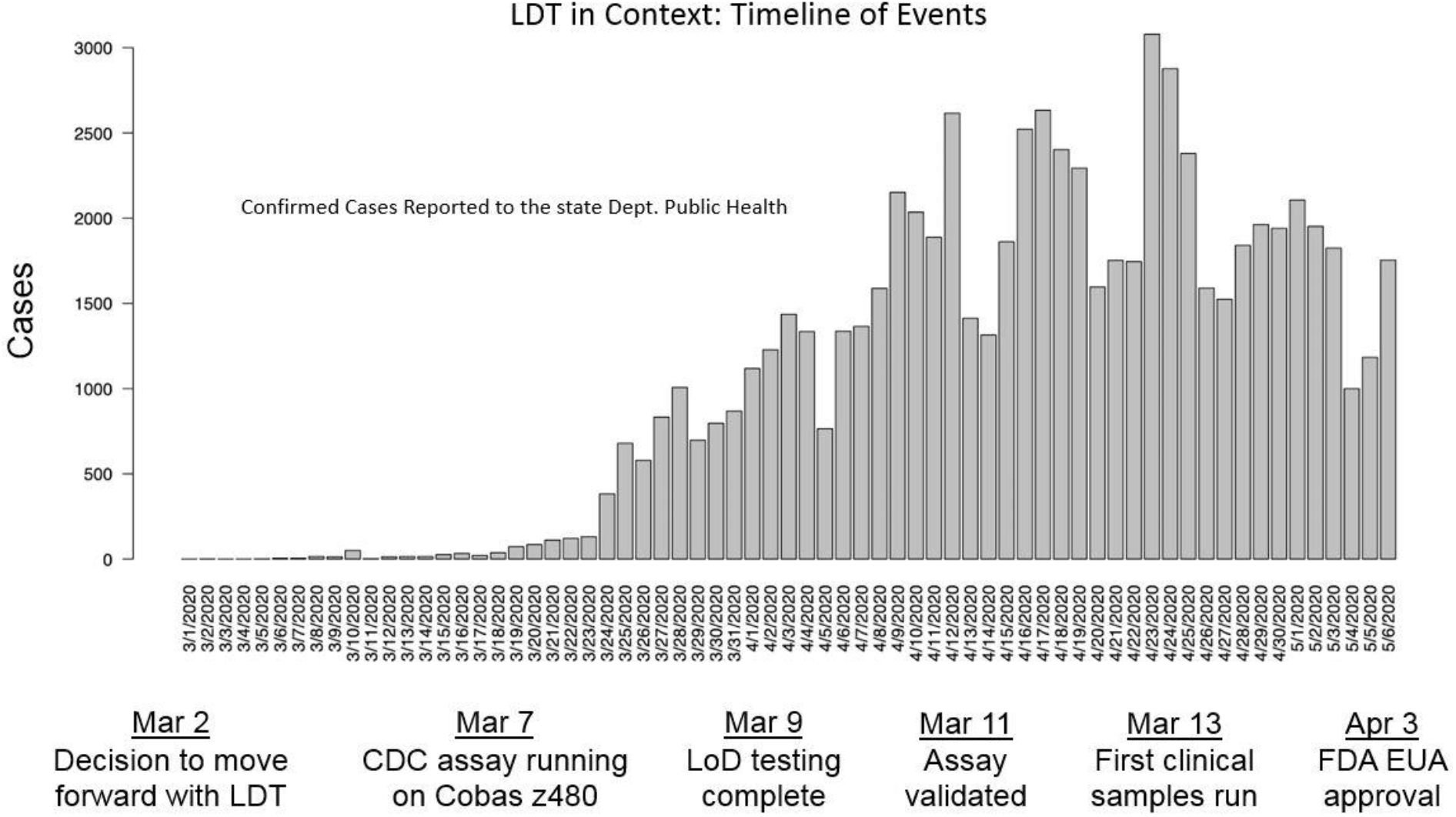
A timeline of events placing the development of the LDT assay in the context of the local epidemic of COVID-19. Histogram showing the showing the local epidemic curve as defined by daily cases reported to the state Public Health Laboratory. Key LDT development milestones are shown below.

We describe here a blueprint for setting up an emergency-use PCR-based LDT. We bolster our suggestions with experience from assay development for SARS-CoV-2, explaining the obstacles and issues that we encountered and the steps we took to address them, in the hope that they will prove useful to other laboratories striving to create and implement their own LDTs. In particular, we detail the necessary steps to address the following six issues: (1) assay design and selection; (2) procurement of personnel, materials, and equipment; (3) laboratory set up and workflows; (4) assessment of the assay’s technical performance; (5) assessment of clinical performance, and (6) additional considerations for clinical deployment. These discrete and easy to follow guidelines encompass the fundamental principles of a PCR diagnostic test and are designed to be applicable regardless of regulatory landscape. While our focus is on the technical aspects of LDT development, we also acknowledge the need for a change in policy to facilitate LDT establishment for emergent pathogens. With the necessary education, resources, and faster moving, readily comprehensible regulatory policies in place, our collective capacity for effective and rapid response efforts will increase significantly, better preparing us for any future outbreaks.

### Step 1: Assay Design and Selection

PCR-based diagnostic assays use short complementary oligonucleotide primers to specifically amplify a specific DNA sequence. PCR assays are often used to directly detect pathogen DNA or, in the case of an RNA virus like SARS-CoV-2, complementary DNA (cDNA) generated from reverse transcription of the RNA genome. Amplification of the specific DNA sequence is then detected in real-time by an intercalating dye, such as SYBR Green, or a fluorescent-labeled probe.

#### Assay Design

The design of the primer and probe sequences is the most critical factor in the development of a highly specific and sensitive assay. Primer sequences must efficiently bind to their target regions and be pathogen-specific to avoid cross-reactivity. In an outbreak scenario, validated primer-probe sets are often available from public health authorities but if not, the following guidelines can be considered. Ideally, primer sequences are placed in highly conserved regions of the genome, as frequent mutation in a primer region can interfere with primer binding and deem an assay useless. In an emerging outbreak, pathogen sequences are scarce and diversity across the genome is poorly understood. However, analyzing related pathogens may help identify regions of the genome that are more likely to be highly conserved, such as structural or housekeeping genes. Another strategy is to target both a region that is conserved amongst very closely related organisms, *e.g*. SARS-like coronaviruses, to ensure sensitivity and a pathogen-specific region to ensure specificity. Potential primer designs should be analyzed to ensure there are no primer-primer interactions or secondary structure inhibition from hairpin structures.^4^ Assays should also incorporate a human gene target that can serve as full process control, both ensuring specimen and extraction quality and acting as a PCR control.

#### Checking for Cross Reactivity

As an important part of assay design, laboratories should assess assay cross-reactivity with closely related pathogens, pathogens that produce similar clinical syndromes or are found in similar anatomic sites, and common commensal organisms using an *in silico* approach. For example, for the SARS-CoV-2 respiratory virus, the FDA required: “At a minimum, an *in silico* analysis of the assay primer and probes compared to common respiratory flora and other viral pathogens… should be performed. FDA defines in silico cross-reactivity as greater than 80% homology between one of the primers/probes and any sequence present in the targeted microorganism.”^5^

To assess primer specificity, we performed BLAST (blastn, using standard parameters) searches for both the N1 and N2 CDC primer and probe sequences against SARS-CoV-2, SARS-CoV-1, MERS-CoV, the four seasonal coronaviridae, 13 other respiratory viruses, 11 respiratory bacterial and mycobacterial pathogens, *Candida albicans*, and *Pneumocystis jirovecii* (see supplemental table). There were no bi-directional primer hits in any organism other than SARS-CoV-2. Based on these data, we consider primers 2019 nCoV_N1 and 2019 nCoV_N2 as highly specific to detect COVID-19. As mentioned above, empirical testing of clinically-validated samples also showed no cross reactivity with human metapneumovirus, influenza A, influenza B, RSV, adenovirus, coronavirus 229E, and parainfluenza (Supplemental Table A1).

We considered several assay designs, including the Drosten assay that was adopted by the World Health Organization (WHO).^6^ However, in order to maintain as much consistency as possible with the US CDC and state public health laboratories, we based our LDT on the existing CDC 2019-nCoV Real-Time RT-PCR Diagnostic panel. We modified the protocol in several key ways and proceeded to validate this amended protocol. We describe these modifications in Step 2 below.

### Step 2: Procuring personnel and materials and optimizing instrumentation

#### Personnel

The development of a new diagnostic test is a personnel-intensive process. During a pandemic, laboratories must still maintain normal operations and finding skilled personnel that can dedicate time to assay development is challenging. If possible, we recommend dedicating one person or a small group to focus solely on assay development. When evaluating personnel options, in addition to the diagnostic laboratory’s clinical personnel, consider partnering with closely affiliated researchers, as we did, for technical development and validation of the LDT. While this may not be feasible for all laboratories, it allowed clinical personnel to maintain essential laboratory operations while pursuing LDT development efforts.

#### Sample Extraction Materials and Methods

In order to perform PCR, laboratories must be able to first extract nucleic acid from a primary biological sample. While there are automated platforms that perform this task at high throughput, they are not widely available. However, manual spin column-based extractions kits, although low-throughput and labor intensive, are widely used, simple, and reliable. Thus, for expediency, we initially used a manual extraction protocol. While many manual extraction kits are available for the isolation of RNA or DNA from a variety of sample sources and volumes, we began our initial validation with the widely-available, FDA-approved QIAamp Viral RNA Mini kit (Qiagen Inc, Germantown MD, USA).

### PCR Reagents

Once a decision on assay selection or design has been made, laboratories must obtain the primers and probes from commercial entities or directly from public health authorities, who often aid in distribution of these key reagents. We ordered a primer and probe kit based on published CDC sequences for N1, N2, and an RNAse P control from Integrated DNA Technologies (10006606). As SARS-CoV-2 is an RNA virus, we needed an RT-PCR master mix. We ordered the Applied Biosystems TaqPath 1-Step RT-qPCR Master Mix from ThermoFisher (Cat. A15300).

#### PCR Instrumentation

PCR reactions require specialized equipment that are capable of thermal cycling and fluorescent signal detection. Most molecular laboratories will have access to a thermocycler. For expediency we began our development with an instrument we had available, a Roche cobas z 480. The use of this instrument required a deviation from the published cycling conditions of the CDC protocol we used as a guide for development. The CDC protocol calls for a 2-minute incubation with uracil-DNA N-glycosylase (UNG) at 25°C, but our instrument’s lowest programmable temperature was 37°C. Since the UNG is active over a relatively broad temperature range, we employed a 5-minute incubation at ambient room temperature (Table 1). Similar adjustments will be necessary as each laboratory optimizes the assay for the equipment and reagents on hand.

**Table 1.** PCR cycling conditions used by the CDC compared to the LDT PCR cycling conditions with a different thermocycler. The CDC used the ABI 7500FastDx, while the LDT protocol used the Roche cobas z480.

|  | **CDC**<br>**cycling conditions**<br>**(ABI 7500FastDx)** |  | **LDT**<br>**cycling conditions**<br>**(Roche cobas z480)** |  |
| --- | --- | --- | --- | --- |
| UNG Incubation | 25°C | 2 mins | 20-23°C | 5 mins |
| RT Incubation | 50°C | 15 mins | 50°C | 15 mins |
| Enzyme Activation | 95°C | 2 mins | 95°C | 2 mins |
| Amplification | 95°C | 3 secs | 95°C | 3 secs |
| (45 cycles) | 55°C | 30 secs | 55°C | 30 secs |

#### Controls

Quality controls are a critical component of molecular assay development but can be challenging to obtain if reference materials are not yet commercially available. Multiple controls are required to comply with laboratory regulations: an internal amplification control for each specimen to demonstrate the absence of a PCR inhibitor, an extraction control to detect problems with nucleic acid extraction, and positive and negative amplification controls for each analyte, as discussed at length elsewhere^7^ and briefly described here. For laboratory-developed tests, the internal amplification control often consists of a human housekeeping gene, *e.g*. ribonuclease P, beta actin, or glyceraldehyde-3-phosphsate dehydrogenase, which is present within the patient specimen at a concentration comparable to the pathogen of interest. Lack of amplification of the internal amplification control indicates the presence of a PCR inhibitor, failure of the nucleic acid extraction step, or insufficient sample collection. In addition, a separate extraction control should be tested with each extraction batch. Ideally, this extraction control is comprised of the whole, inactivated pathogen spiked into a clinical matrix at a low concentration to mimic a low positive sample and improve the likelihood of detecting suboptimal extraction performance. Finally, positive and negative amplification controls must be tested with each batch to verify successful amplification conditions and the absence of reagent or specimen contamination, respectively. The positive control can be positive patient specimens, pathogen spiked into a clinical matrix, purified pathogen nucleic acid, or synthetic gene targets. The negative control can be known negative patient specimens or nuclease-free water.

Early in the SARS-CoV-2 pandemic, we did not have a readily available source of SARS-CoV-2 viral RNA control material or positive patient specimens. For a positive control, we initially used in vitro transcribed (IVT) RNA from the full-length SARS-CoV-2 N gene (GenBank accession: MN908947.2), the region targeted by the CDC assay (gifted from Sherlock Biosciences). We stored 50 μL aliquots of the 1 nM IVT RNA in single-use aliquots at −80 °C to avoid RNA degradation from multiple freeze-thaw cycles. If RNA is unavailable, a DNA synthetic positive control is a possible alternative. Our negative control was pooled negative sample matrix, comprised of nasopharyngeal specimens collected a year prior to the emergence of SARS-CoV-2, which was carried through the extraction and amplification process. This negative control also served as an extraction control in lieu of positive patient specimens. Once positive patient specimens became available, we developed a new extraction control comprised of a high-titer patient specimen that we diluted into a large volume of pooled negative sample matrix and froze in aliquots for storage at −80 °C.

### Step 3: Space Planning for a Clean Molecular Workflow and Laboratory Safety

Contamination is a particular concern in a clinical molecular diagnostics laboratory, where spurious amplification of nucleic acids can lead to false positives results.^8–10^ To minimize this risk, we established a unidirectional workflow for each PCR step carried out in a workspace dedicated to that step. All reagent preparation (master mix preparation, aliquoting of extraction kit reagents, etc.) was done in a clean area, which we designate as Area 1, with a strict policy that no target material, sample material, or personnel who had handled the aforementioned material that same day could enter the area. All RNA extractions and addition of RNA to the PCR plate were done in a separate physical space, Area 2. In addition, a third physical space, Area 3, was designated for amplification of nucleic acids, to keep post-PCR amplicons physically separated from unamplified clinical specimens. If discrete areas are not available, it is best to pay extra attention to the directionality of the daily workflow and try to separate each step as much as physically possible.

We also employed specific laboratory practices to minimize contamination and RNA degradation. For example, because high concentration synthetic genetic material is often a source of contamination and must be handled carefully, we diluted all stocks to 1E+5 copies/uL or lower in molecular-grade Tris-EDTA buffer before introducing them to the working area. RNA is also sensitive to degradation by RNases. To help prevent degradation and contamination, we employed ultra-clean laboratory practices, ensured proper temperature and pH, used EDTA and carrier RNA when appropriate, and used molecular grade reagents and consumables such as aerosol tips. Further, we consistently avoided freeze-thaw cycles and wiped all surfaces and equipment used for RNA work with RNAse Zap and freshly prepared 10% bleach.

In addition, it is critical to ensure the safety of laboratory staff who are working with a novel pathogen when limited information is available on transmission risk. The precise biosafety controls depend on the biosafety level (BSL) categorization assigned to the pathogen and may be influenced by the level of risk perceived by laboratorians. For example, the WHO and CDC recommend performing RNA extraction for SARS-CoV-2 testing in a biosafety cabinet contained within a BSL-2 facility with standard precautions and without prior heat inactivation of the sample.^11^ However, especially early in the pandemic, many laboratorians preferred to don additional personal protective equipment and perform heat inactivation for peace of mind. Collecting specimens directly into molecular transport media to inactivate microbes and stabilize nucleic acid provides an additional safety control, though it comes at increased cost and requires additional clinical validation.

### Step 4: Establishing the Analytic Sensitivity

The first step of the development and characterization of any laboratory developed test is to determine the parameters of the technical performance of the assay. First and foremost, laboratories must determine the analytic sensitivity of the assay, which is commonly referred to as the limit of detection (LoD). While the limit of detection is influenced by many factors, including but not limited to, sample type, extraction efficiency, input volume, and assay design, PCR’s theoretical sensitivity is as low as 1 target copy per reaction. To efficiently determine the LoD, the LoD can first be approximated by testing a wide range of concentrations and then confirmed with many replicates at a single concentration.

#### A) Initial LoD assessment

During initial assay development, we did not have access to SARS-CoV-2-positive patient samples or full-length SARS-CoV-2 RNA. Instead, we first roughly estimated our LoD with a pure DNA template input, followed by a series of contrived positive samples spanning 1E+3 to 1E+0 copies per μL that were tested in triplicate (Table 2). For the DNA template input, we tested both custom N gene DNA gBlocks (double-stranded DNA fragments) and DNA plasmid controls of the entire SARS-CoV-2 N gene (Integrated DNA Technologies, Coralville, IA). For the contrived samples, we spiked IVT RNA of the SARS-CoV-2 N gene into pooled SARS-CoV-2-negative nasopharyngeal (NP) specimens collected in universal transport media (UTM) (Copan, UTM-RT), which was the preferred specimen type for testing. Notably, unprotected IVT RNA is rapidly degraded by endogenous RNases if added directly to the specimen. Rather, RNases must be inactivated prior to RNA spike-in, for example by mixing the clinical sample matrix with a guanidinium thiocyanate-containing buffer (e.g. Qiagen’s Buffer AVL) that is used in downstream RNA extraction. The IVT RNA can then be added to the specimen and carried through the subsequent RNA extraction steps. If RNA is not available, laboratories will have to assess RNA extraction and cDNA detection steps independently.

**Table 2.** Initial limit of detection results for LDT qPCR assay.

| **2019-nCoV_N1** |  |  |  |  |  |  |  |
| --- | --- | --- | --- | --- | --- | --- | --- |
| **Genomes/μL** | 0 | 0.37 | 1.11 | 3.3 | 10 | 100 | 1000 |
| **% positive (out of 3)** | - | 0% | 67% | 100% | 100% | 100% | 100% |
| **Mean Ct (SD)** | - | - | - | 34.8 (0.35) | 33.14 (0.13) | 29.89 (0.15) | 25.99 (0.1) |
| **2019-nCoV_N2** |  |  |  |  |  |  |  |
| **Genomes/μL** | 0 | 0.37 | 1.11 | 3.3 | 10 | 100 | 1000 |
| **% positive (out of 3)** | - | 33% | 67% | 100% | 100% | 100% | 100% |
| **Mean Ct (SD)** | - | - | - | 40 (0) | 37.98 (0.51) | 34.17 (0.31) | 29.66 (0.21) |
| **RNase P** |  |  |  |  |  |  |  |
| **Genomes/μL** | 0 | 0.37 | 1.11 | 3.3 | 10 | 100 | 1000 |
| **% positive (out of 3)** | 67% | 100% | 100% | 100% | 100% | 100% | 100% |
| **Mean Ct (SD)** | 28.01 (0.02) | 28 (0.02) | 28 (0.02) | 28 (0.13) | 28 (0.04) | 28.01 (0.05) | 27.96 (0.2) |

#### B) LoD confirmation

Under emergency circumstances, the FDA has allowed laboratories to confirm the LoD while simultaneously assessing assay precision by demonstrating a ≥95% detection rate with 20 replicates. The LoD confirmation experiments should be performed using the most complex specimen type that will be tested with the assay. Ideally, the 20 replicates would be performed with unique specimens rather than a pooled matrix to increase robustness to specimen differences and over several days with different operators.

We chose a target concentration of 5 copies/μL, based on our earlier estimate that the LoD was between 10 and 3.3 copies/μl sample input. A total of 19 out of 20 samples tested positive at 5 copies/μL (20/20 for N1 set, 19/20 for N2 set). The 5 copies/uL LoD is slightly higher than the reported LoD for the CDC assay (1 copy/μL). Possible explanations include: (1) matrix differences (the CDC used a A549 lung adenocarcinoma cell line in UTM, compared to our use of pooled clinical samples); (2) possible quantitation differences and dilutional error stacking associated with our custom-made transcribed RNA stock; or (3) differences in PCR instrumentation and cycling speeds. We observed a slightly lower efficiency of the N2-detection reagents compared to the efficiency reported in the CDC package insert. We found an approximately four-cycle lag with the N2 primer set when compared to the N1 set, while the results from the CDC indicated that the difference should be closer to one cycle. Further work at a collaborating institution later revealed that the N2-set performance could be rescued by extending the melt time to 5-10 seconds on instruments not capable of fast cycling. Nevertheless, we proceeded under the expectation that the process and LoD differences would not be clinically significant.

Before beginning our clinical validation, we also defined the criteria for a positive result. We considered two major factors: cycle threshold setting and Ct-value validity range. Based on a review of all data from our analytical validation runs, we decided on the following settings for our qPCR instrument: (1) the noise band would be set manually above the highest negative sample in the run; (2) for a valid positive result, the cycle threshold values from both the N1 and N2 targets must be below 42.5; and (3) for a valid negative result, the human RNase P control target must also amplify successfully, with a Ct<35 signifying adequate sample collection, a more stringent cutoff than the CDC’s protocol (RNase P Ct less than 40).^12^ Although these settings differed slightly from those used by the CDC, the performance data was sufficiently reproducible to allow these accommodations for our instrument-related differences.

### Step 5: Determining Clinical Accuracy

Having determined the LoD and defined interpretative criteria, we next determined the accuracy of our LDT on primary clinical specimens. Clinical specimens used in the validation study should be selected to challenge the assay’s sensitivity while confirming its specificity. Testing is performed in a randomized and blinded manner. The number of positive and negative specimens to test is at the laboratory’s discretion, but the FDA guidance for emergency-use SARS-CoV-2 assays required at least 30 reactive and 30 non-reactive specimens.

If known positive specimens are not available, the FDA has allowed contrived reactive specimens to be used: “Contrived reactive specimens can be created by spiking RNA or inactivated virus into leftover individual clinical specimens representing unique patients; the majority of these specimens can be leftover respiratory specimens such as NP swabs, sputum, etc. Twenty of the contrived clinical specimens should be spiked at a concentration of 1x-2x LoD, with the remainder of specimens spanning the assay testing range.”^13^ Following this guidance, we selected thirty unique clinical nasopharyngeal specimens collected a year prior to the emergence of SARS-CoV-2: 15 were negative for all clinically-tested respiratory viruses and 15 that had tested positive for either influenza A, influenza B, respiratory syncytial virus, human metapneumovirus, parainfluenza virus, adenovirus, and/or coronavirus 229E (Supplemental Table A2). The latter set allowed us to evaluate potential cross-reactivity and PCR interference from organisms likely to be encountered in respiratory samples. Each specimen was divided into two aliquots: one aliquot was spiked with SARS-CoV-2 transcribed RNA as a contrived reactive specimen, and the second was spiked with an equivalent amount of Qiagen AVE buffer (the diluent used in our earlier RNA dilution series) as a negative specimen. For our contrived reactive specimens, we spiked 20 samples with 10 copies/μL of IVT N gene RNA (~2x LoD), enabling us to assess assay performance near LoD; 15 of these were known to contain other respiratory viruses, allowing us to further assess interference. We spiked the remaining 10 contrived positive samples with varying amounts of RNA to represent a range of viral burdens from 100 - 10,000 copies/μL (Supplemental Table A3). We then tested these sixty specimens on our assay (Figure 2).

**Figure 2:**
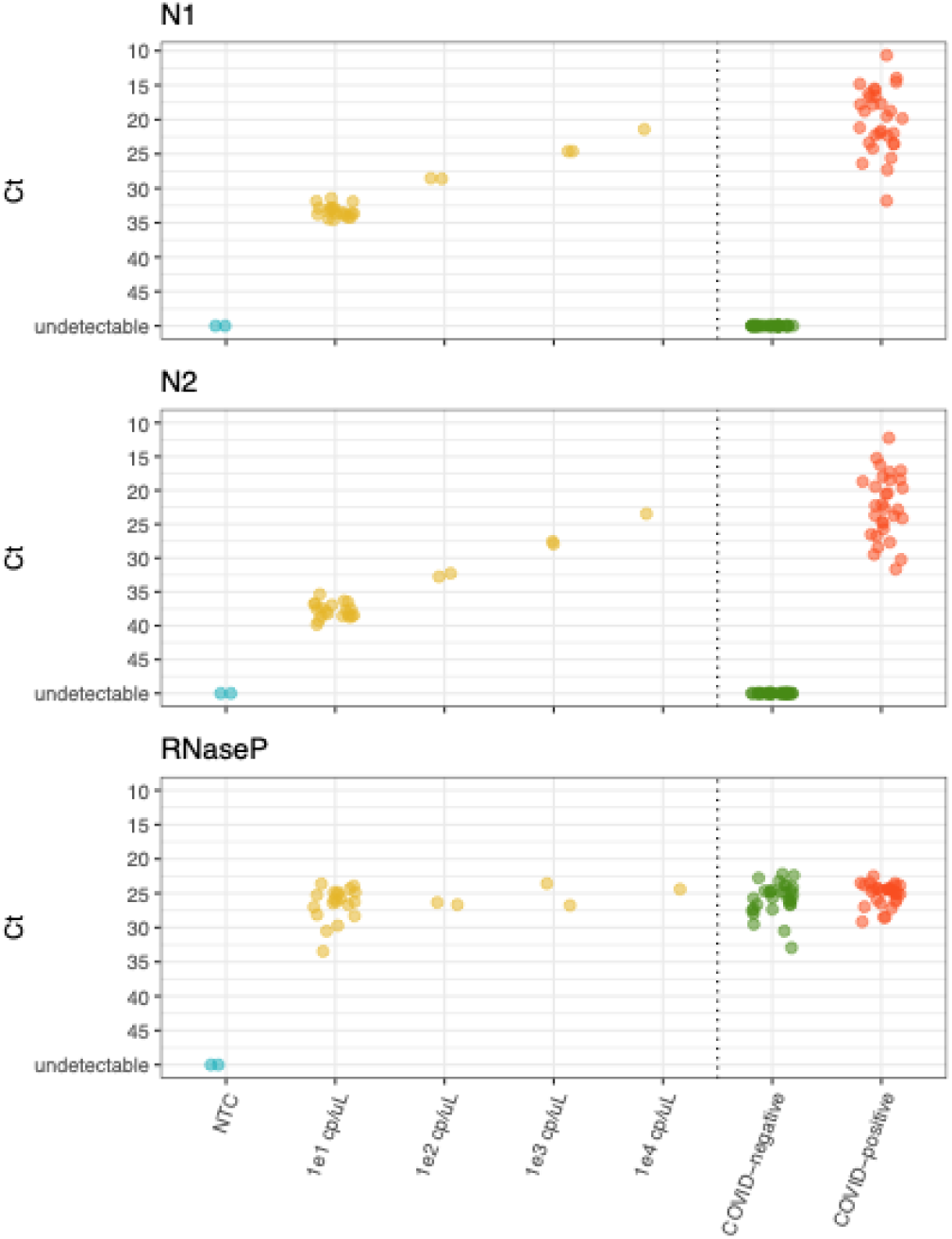
Clinical performance assessment in COVID-positive specimens, contrived positive specimens, and negative controls. Ct (cycle threshold) values for N1, N2, and RNAse P primer-probe sets are shown. A negative control, NTC (no template control), is shown in blue. Contrived positive NP samples are represented in yellow, at four different spike-in concentrations (from 10 to 10000 copies/μL). Clinical samples, at the right of the figure, comprise known COVID-negative samples (green) and COVID-positive samples (red). N1 and N2 represent reactions with SARS-CoV-2 specific primer pairs, with primer sequences consistent with those published by the CDC. RNase P primers amplify human RNA and thus these reactions serve as positive controls to ensure that these specimens do not contain significant PCR inhibitors and have adequate sample quality.

While performing our clinical validation, we began to see COVID-19 cases and were able to extend the validation to include 30 additional samples that tested positive for SARS-CoV-2 at the State Public Health Laboratory. These samples were tested directly and not diluted to near-LoD concentrations. The results were uniformly concordant with those from the State laboratory (Supplemental Table A4).

### Step 6: Additional Considerations for Clinical Deployment

Once a laboratory has confidence in their assay and can demonstrate proper regulatory compliance, it will need to begin a transition to clinical deployment. This will vary widely from laboratory to laboratory, as hospital-based laboratories, reference laboratories, and other testing centers will have different considerations to take into account. In addition to transitioning to clinical use, laboratories should look for ways to scale up testing capacity as the need for testing increases. We describe some additional considerations that proved critical as our laboratory began to transition from development to clinical deployment and scale up our processes.

The process of bringing a new LDT into use in a clinical laboratory usually takes months of training and planning; however, the ongoing pandemic required an accelerated timeline. In an effort to bring the assay to clinical use as quickly as possible, we took the following steps to smooth the transition from laboratory testing to clinical deployment. We performed as much validation work as possible in the clinical space with clinical laboratory personnel. We carried out our clean area work, such as reagent preparation and plate stamping, in the clinical laboratory spaces so that we could easily transfer those steps of the process to the clinical laboratory staff. (We did, however, keep our high-copy SARS-CoV-2 control nucleic acid out of the clinical setting to minimize contamination risk.) Towards the end of the validation process, clinical technologists observed the validation team and then worked side-by-side to complete proficiency training. Another important piece of clinical validity of an assay is continued quality assurance, assessed using a positive control and, eventually, external proficiency samples. Positive control material will likely not be commercially available early in an outbreak but laboratories can make their own using synthetic targets or real patient samples, if available. Ideally, the positive control also assesses assay performance near the LoD, so we prepared a large volume of sample spiked at a concentration of ~2-5X LoD. The steps we took to accelerate personnel training and clinical deployment of the assay allowed us to begin clinical testing quickly after validation work was complete.

Once an assay has been established, laboratories should assess their ability to scale-up testing if there are adequate resources to do so. It is important to note that when bringing new equipment or reagents online, repeating the entire validation is not necessary. While regulations may vary, we recommend that laboratories perform a bridging study to determine equivalency in performance. As a case example, after our laboratory found that manual extraction kits would not provide a sustainable solution to meet the testing demand, we were able to purchase an automated high-throughput extraction platform (Roche MagnaPure 24) and performed a bridging study to demonstrate equivalency with manual extraction and bring that equipment online.

## DISCUSSION

In the early stages of a pandemic, rapid and accurate testing forms the centerpiece of a coordinated response, directly influencing both public health efforts and patient care. When the COVID-19 pandemic arrived in the U.S., it was critically important to ramp up high-quality testing as quickly as possible. In partnership with collaborating institutions and the state DPH, our team became one of the first hospitals in the country to have an FDA EUA-approved laboratory-developed test.^14^ Our LDT filled a critical gap for 2-3 weeks when there was no alternative diagnostic test for COVID-19. During this time, the LDT facilitated enrollment of patients into key clinical trials and helped guide care of the most critically ill patients due to its faster turn-around time than send-out testing options.^15,16^ After this initial period of turmoil, the LDT became a crucial resource to validate higher-throughput platforms and new specimen-types.

We hope that sharing our experience can provide a useful roadmap for other laboratories attempting to set up their own assays, both for continued SARS-CoV-2 testing and for future infectious disease outbreaks. Our approaches are generalizable to almost any clinical laboratory hoping to set up a molecular-based LDT for pathogen detection. While new molecular technologies such as CRISPR-based detection are in development, the widespread capacity and versatility of PCR-based diagnostics allow this type of diagnostic test to be readily, widely, and cheaply deployed.

When pursuing an LDT, laboratories will first need to select an assay. Laboratories may choose to design their own assay or adopt an assay already in use in their state or country. Critically, assay design and selection should be dictated by the resources a laboratory has on hand to allow for the fastest possible development and deployment. Assay validation should proceed in a stepwise manner, first establishing the assay’s limit of detection, and then proceeding to a clinical validation step undertaken with the most challenging sample type that the laboratory is planning to test diagnostically. Finally, when transitioning the LDT into clinical use, laboratories should take special consideration to uphold quality assurance with constant use of reliable positive and negative controls. If resources are available, testing capacity should be scaled up to accommodate the need for high volume testing. Throughout the development and deployment of our LDT, we relied on new and existing collaborations for success. We also remained creative and resourceful in our usage of instrumentation and reagents to achieve our goal of validating the assay as quickly and safely as possible, and then moving it into clinical practice.

As a concrete example of the widely-deployable nature of these PCR-based LDTs, we worked with both local and international partners to rapidly set up COVID-19 testing early in the outbreak in a number of different settings around the world. Our partners in Nigeria, Sierra Leone, and Senegal had established LDTs by February, within days of the public release of SARS-CoV-2 genome sequence data. We also consulted for other U.S. based laboratories as they set up LDTs.

Now more than ever, clinical laboratories throughout the world desperately need expanded access to molecular diagnostic tests that can be performed at scale. Given the challenges in expanding access to existing commercially available high-throughput molecular diagnostic platforms, we hope other laboratories can rapidly respond if necessary, by standing up molecular diagnostics independently, even in the midst of a global pandemic. Ultimately, our success in this and future pandemics will require a major shift in both policy and practice, to empower more actors to build LDTs that produce accurate results early in an outbreak and conduct testing wherever needed. We hope that the tools and techniques we describe here, based on our experience, will help facilitate a collective increase in capacity, enabling deployment of LDTs within just days of novel pathogen detection.

## Data Availability

All of the data is either shared in the manuscript or can be made available on request.

## Funding and Acknowledgements

Parts of this work were supported through a grant from the Centers for Disease Control and Prevention [U01 CK000490]; as well as a grant from the National Institutes of Health/National Institute of Allergy and Infectious Diseases [NIH/NIAID 2U19AI110818-06].

## Disclaimers (Conflicts of Interest)

Pardis Sabeti is a founder and shareholder of Sherlock Biosciences, and is both on the Board and serves as shareholder of the Danaher Corporation. Jacob Lemieux is a consultant for Sherlock Biosciences. Melis N. Anahtar is a co-founder, equity holder, and consultant for Day Zero Diagnostics. Pardis Sabeti, Edward T. Ryan, and Sarah E. Turbett have received CDC funding for this work and other COVID-related work. John Branda has received grant support from Zeus Scientific, bioMerieux, Immunetics, the Bay Area Lyme Foundation and the Lyme Disease Biobank Foundation for work unrelated to this study, and has been a consultant for T2 Biosystems, DiaSorin and Roche Diagnostics.

## Supplemental Materials

**Supplemental Table A1:**
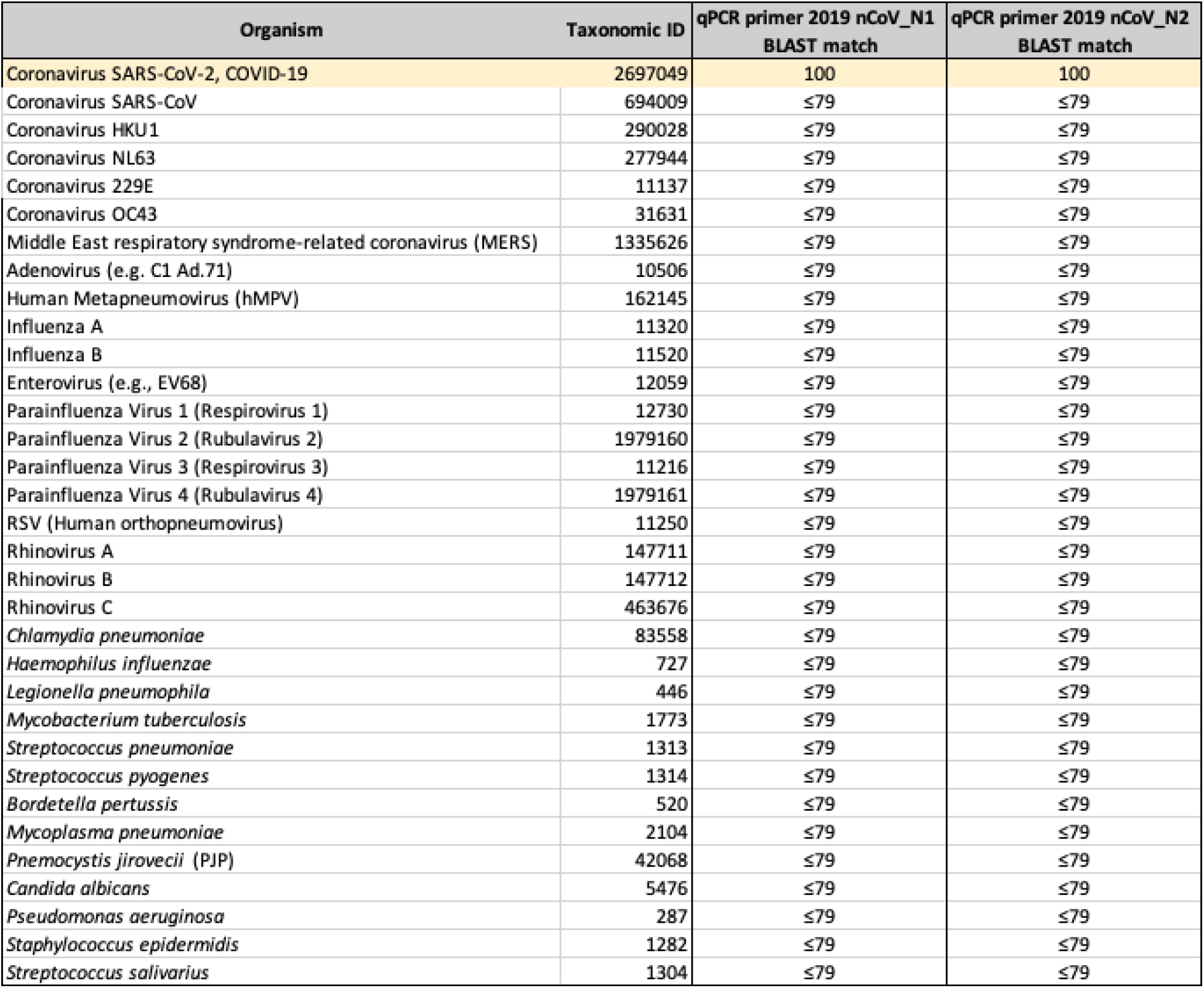
In silico cross-reactivity assessment of N1 and N2 primer pairs across a variety of common respiratory pathogens.

| Organism | Taxonomic ID | qPCR primer 2019 nCoV_N1<br>BLAST match | qPCR primer 2019 nCoV_N2<br>BLAST match |
| --- | --- | --- | --- |
| Coronavirus SARS-CoV-2, COVID-19 | 2697049 | 100 | 100 |
| Coronavirus SARS-CoV | 694009 | ≤79 | ≤79 |
| Coronavirus HKU1 | 290028 | ≤79 | ≤79 |
| Coronavirus NL63 | 277944 | ≤79 | ≤79 |
| Coronavirus 229E | 11137 | ≤79 | ≤79 |
| Coronavirus OC43 | 31631 | ≤79 | ≤79 |
| Middle East respiratory syndrome-related coronavirus (MERS) | 1335626 | ≤79 | ≤79 |
| Adenovirus (e.g. C1 Ad.71) | 10506 | ≤79 | ≤79 |
| Human Metapneumovirus (hMPV) | 162145 | ≤79 | ≤79 |
| Influenza A | 11320 | ≤79 | ≤79 |
| Influenza B | 11520 | ≤79 | ≤79 |
| Enterovirus (e.g., EV68) | 12059 | ≤79 | ≤79 |
| Parainfluenza Virus 1 (Respirovirus 1) | 12730 | ≤79 | ≤79 |
| Parainfluenza Virus 2 (Rubulavirus 2) | 1979160 | ≤79 | ≤79 |
| Parainfluenza Virus 3 (Respirovirus 3) | 11216 | ≤79 | ≤79 |
| Parainfluenza Virus 4 (Rubulavirus 4) | 1979161 | ≤79 | ≤79 |
| RSV (Human orthopneumovirus) | 11250 | ≤79 | ≤79 |
| Rhinovirus A | 147711 | ≤79 | ≤79 |
| Rhinovirus B | 147712 | ≤79 | ≤79 |
| Rhinovirus C | 463676 | ≤79 | ≤79 |
| *Chlamydia pneumoniae* | 83558 | ≤79 | ≤79 |
| *Haemophilus influenzae* | 727 | ≤79 | ≤79 |
| *Legionella pneumophila* | 446 | ≤79 | ≤79 |
| *Mycobacterium tuberculosis* | 1773 | ≤79 | ≤79 |
| *Streptococcus pneumoniae* | 1313 | ≤79 | ≤79 |
| *Streptococcus pyogenes* | 1314 | ≤79 | ≤79 |
| *Bordetella pertussis* | 520 | ≤79 | ≤79 |
| *Mycoplasma pneumoniae* | 2104 | ≤79 | ≤79 |
| *Pneumocystis jirovecii* (PJP) | 42068 | ≤79 | ≤79 |
| *Candida albicans* | 5476 | ≤79 | ≤79 |
| *Pseudomonas aeruginosa* | 287 | ≤79 | ≤79 |
| *Staphylococcus epidermidis* | 1282 | ≤79 | ≤79 |
| *Streptococcus salivarius* | 1304 | ≤79 | ≤79 |

**Supplemental Table A2:** Assay performance in COVID-negative samples and in-vitro assessment of cross-reactivity.

| COVID-negative Sample Number | Clinical results for additional respiratory pathogens | 2019 nCov_N1 Ct | 2019 nCov_N2 Ct | RNAse P Ct | Result | MA DPH Result | Concordance |
| --- | --- | --- | --- | --- | --- | --- | --- |
| COVID-negative 1 | Negative<sup>1</sup> | — | — | 24.67 | Negative |  | Yes |
| COVID-negative 2 | Negative<sup>1</sup> | — | — | 22.77 | Negative |  | Yes |
| COVID-negative 3 | Negative<sup>1</sup> | 34.93<sup>3</sup> | — | 25.33 | Negative |  | Yes |
| COVID-negative 4 | Negative<sup>1</sup> | — | — | 25.23 | Negative |  | Yes |
| COVID-negative 5 | Negative<sup>1</sup> | — | — | 26.64 | Negative |  | Yes |
| COVID-negative 6 | Negative<sup>1</sup> | — | — | 24.57 | Negative |  | Yes |
| COVID-negative 7 | Negative<sup>1</sup> | — | — | 26.68 | Negative |  | Yes |
| COVID-negative 8 | Negative<sup>1</sup> | — | — | 26.64 | Negative |  | Yes |
| COVID-negative 9 | Negative<sup>1</sup> | — | — | 22.21 | Negative |  | Yes |
| COVID-negative 10 | Negative<sup>1</sup> | — | — | 24.81 | Negative |  | Yes |
| COVID-negative 11 | Negative<sup>1</sup> | — | — | 25.68 | Negative |  | Yes |
| COVID-negative 12 | Negative<sup>1</sup> | — | — | 23.85 | Negative |  | Yes |
| COVID-negative 13 | Negative<sup>1</sup> | — | — | 24.64 | Negative |  | Yes |
| COVID-negative 14 | Negative<sup>1</sup> | — | — | 30.5 | Negative |  | Yes |
| COVID-negative 15 | Negative<sup>1</sup> | — | — | 25.75 | Negative |  | Yes |
| COVID-negative 16 | Human Metapneumovirus | — | — | 26.13 | Negative |  | Yes |
| COVID-negative 17 | Adenovirus, Coronavirus 229E | — | — | 27.34 | Negative |  | Yes |
| COVID-negative 18 | Adenovirus | — | — | 25.96 | Negative |  | Yes |
| COVID-negative 19 | Adenovirus | — | — | 27.3 | Negative |  | Yes |
| COVID-negative 20 | Parainfluenza Virus 3 | — | — | 27.99 | Negative |  | Yes |
| COVID-negative 21 | Parainfluenza Virus 3 | — | — | 23.84 | Negative |  | Yes |
| COVID-negative 22 | Parainfluenza Virus 2 | — | — | 32.95 | Negative |  | Yes |
| COVID-negative 23 | Respiratory Syncytial Virus | — | — | 24.84 | Negative |  | Yes |
| COVID-negative 24 | Respiratory Syncytial Virus | — | — | 23.22 | Negative |  | Yes |
| COVID-negative 25 | Influenza B | — | — | 32.95 | Negative |  | Yes |
| COVID-negative 26 | Influenza B | — | — | 24.12 | Negative |  | Yes |
| COVID-negative 27 | influenza A/H1-2009 | — | — | 22.41 | Negative |  | Yes |
| COVID-negative 28 | influenza A/H1-2010 | — | — | 24.93 | Negative |  | Yes |
| COVID-negative 29 | influenza A/H1-2011 | — | — | 29.53 | Negative |  | Yes |
| COVID-negative 30 | Negative<sup>2</sup> | — | — | 23.99 | Negative | Negative<sup>4</sup> | Yes |

<sup>1</sup> Negative by a multiplexed PCR for influenza A, influenza B, and RSV (Cepheid) and direct immunofluorescence antigen testing for adenovirus, human metapneumovirus, and parainfluenza types 1-3 (Light Diagnostics).
<sup>2</sup> Negative by a multiplexed PCR for influenza A, influenza B, and RSV (Cepheid).
<sup>3</sup> An aliquot of the NP sample was extracted and run on 3/9 Plate 1, yielding an N1 Ct of 34.93, no amplification with N2, and RNAseP Ct of 25.33. The same extracted RNA was re-run in duplicate on 3/12 Plate 1, and one reaction also gave an N1 Ct of 34.88 and the second reaction did not have amplification of N1. Thus, on 3/13, a new aliquot of the original NP sample was obtained, re-extracted, and re-PCR'd, which had no amplification of N1 or N2, and an RNAse P Ct of 27.52. Thus, the sample was determined to be negative.
<sup>4</sup> A dedicated NP sample and OP sample were sent to the MA State Lab for testing; the NP tested positive with an N1 Ct of 32.05 and N2 Ct of 31.40, and the OP swab was negative. A different NP swab collected on the same day (for flu/RSV testing) was tested by the MGH COVID-19 assay and did not amplify with the N1 or N2 primer/probe sets, while the RNAse P Ct was 23.99. The same extracted RNA was re-tested at MGH and again did not amplify with N1 or N2, while the RNAse P Ct was 24.79. Finally, we sent the MA State Lab the MGH NP sample and they found that it was negative for COVID-19. Thus, we believe there was a pre-analytic error, e.g. sample mislabeling, that led to an erroneous initial result of COVID-19 positivity.

**Supplemental Table A3:**
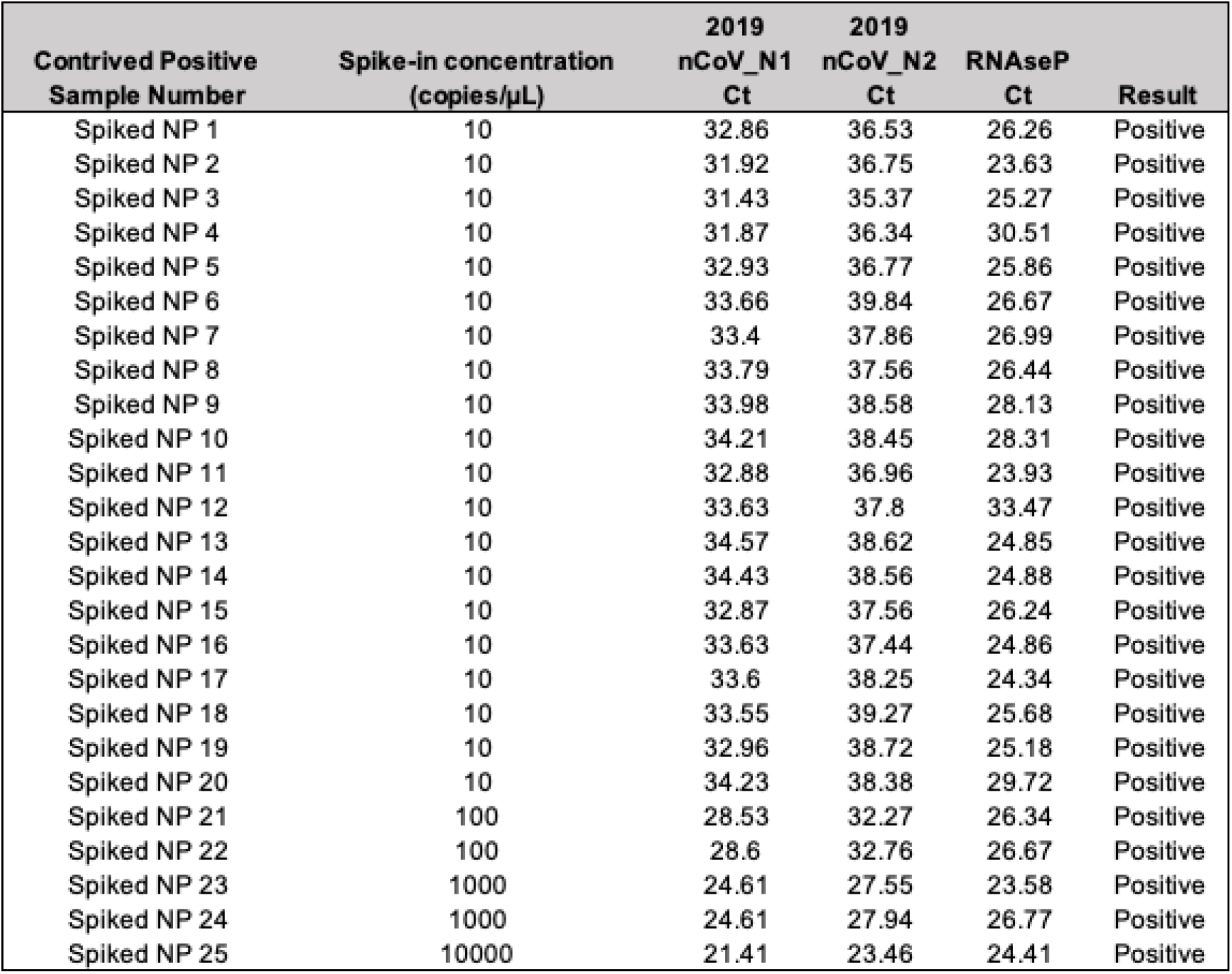
Assay performance in contrived COVID-positive samples.

**Supplemental Table A4:** Assay performance in COVID-positive clinical samples.

| COVID-positive Sample Number | Clinical results for additional respiratory pathogens | 2019 nCov_N1 Ct | 2019 nCov_N2 Ct | RNAse P Ct | Result | MA DPH Result | Concordance |
| --- | --- | --- | --- | --- | --- | --- | --- |
| COVID-positive 1 | Negative<sup>1</sup> | 31.8 | 22.84 | 24.37 | Positive | Positive | Yes |
| COVID-positive 2 | Negative<sup>1</sup> | 16.59 | 18.69 | 24.73 | Positive | Positive | Yes |
| COVID-positive 3 | Negative<sup>1</sup> | 17.8 | 20.45 | 23.43 | Positive | Positive | Yes |
| COVID-positive 4 | Negative<sup>1</sup> | 13.96 | 15.26 | 23.54 | Positive | Positive | Yes |
| COVID-positive 5 | Negative<sup>1</sup> | 17.84 | 19.66 | 24.27 | Positive | Positive | Yes |
| COVID-positive 6 | Negative<sup>1</sup> | 16.25 | 18.4 | 29.15 | Positive | Positive | Yes |
| COVID-positive 7 | Negative<sup>1</sup> | 23.57 | 26.74 | 26.24 | Positive | Positive | Yes |
| COVID-positive 8 | Negative<sup>1</sup> | 21.97 | 24.12 | 28.5 | Positive | Positive | Yes |
| COVID-positive 9 | Negative<sup>1</sup> | 25.6 | 29.49 | 23.5 | Positive | Positive | Yes |
| COVID-positive 10 | Negative<sup>1</sup> | 10.64 | 12.25 | 24.46 | Positive | Positive | Yes |
| COVID-positive 11 | Negative<sup>1</sup> | 19.83 | 22.63 | 24.93 | Positive | Positive | Yes |
| COVID-positive 12 | Negative<sup>1</sup> | 15.5 | 17.3 | 24.42 | Positive | Positive | Yes |
| COVID-positive 13 | Negative<sup>1</sup> | 21.17 | 23.73 | 23.9 | Positive | Positive | Yes |
| COVID-positive 14 | Negative<sup>1</sup> | 18.76 | 20.56 | 24.67 | Positive | Positive | Yes |
| COVID-positive 15 | Negative<sup>1</sup> | 27.31 | 31.65 | 24.92 | Positive | Positive | Yes |
| COVID-positive 16 | Negative<sup>1</sup> | 18.76 | 22.1 | 26.97 | Positive | Positive | Yes |
| COVID-positive 17 | Negative<sup>1</sup> | 23.5 | 28.41 | 27.21 | Positive | Positive | Yes |
| COVID-positive 18 | Negative<sup>1</sup> | 22.36 | 25.72 | 23.85 | Positive | Positive | Yes |
| COVID-positive 19 | Negative<sup>1</sup> | 24.17 | 27.67 | 25.92 | Positive | Positive | Yes |
| COVID-positive 20 | Negative<sup>1</sup> | 23.39 | 26.51 | 26.51 | Positive | Positive | Yes |
| COVID-positive 21 | Negative<sup>1</sup> | 21.62 | 23.7 | 28.52 | Positive | Positive | Yes |
| COVID-positive 22 | Negative<sup>1</sup> | 26.39 | 30.25 | 25.13 | Positive | Positive | Yes |
| COVID-positive 23 | Negative<sup>1</sup> | 15.7 | 17.98 | 25.93 | Positive | Positive | Yes |
| COVID-positive 24 | Negative<sup>1</sup> | 14.8 | 16.27 | 24.27 | Positive | Positive | Yes |
| COVID-positive 25 | Negative<sup>1</sup> | 17.7 | 19.46 | 24.46 | Positive | Positive | Yes |
| COVID-positive 26 | Negative<sup>1</sup> | 16.73 | 18.48 | 23.8 | Positive | Positive | Yes |
| COVID-positive 27 | Negative<sup>1</sup> | 22.26 | 24.61 | 32.95 | Positive | Positive | Yes |
| COVID-positive 28 | Negative<sup>1</sup> | 21.98 | 24.89 | 24.39 | Positive | Positive | Yes |
| COVID-positive 29 | Negative<sup>1</sup> | 14.55 | 17.08 | 25.11 | Positive | Positive | Yes |
| COVID-positive 30 | Negative<sup>1</sup> | 19.51 | 22.25 | 22.51 | Positive | Positive | Yes |

<sup>1</sup> Negative by a multiplexed PCR for influenza A, influenza B, and RSV (Cepheid).

